# Using multi-trait polygenic scores to predict lithium responsiveness in patients with bipolar disorder

**DOI:** 10.64898/2026.01.01.25343187

**Authors:** Nigussie T. Sharew, Scott R. Clark, Simon Hartmann, Sergi Papiol, Urs Heilbronner, Franziska Degenhardt, Janice M. Fullerton, Liping Hou, Tatyana Shekhtman, Mazda Adli, Nirmala Akula, Kazufumi Akiyama, Raffaella Ardau, Bárbara Arias, Fasil Tekola-Ayele, Carla Gallo, Roland Hasler, Hélène Richard-Lepouriel, Nader Perroud, Lena Backlund, Abesh Kumar Bhattacharjee, Frank Bellivier, Antonio Benabarre, Susanne Bengesser, Joanna M. Biernacka, Armin Birner, Cynthia Marie-Claire, Pablo Cervantes, Hsi-Chung Chen, Caterina Chillotti, Sven Cichon, Cristiana Cruceanu, Piotr M. Czerski, Nina Dalkner, Maria Del Zompo, J. Raymond DePaulo, Bruno Étain, Stephane Jamain, Peter Falkai, Andreas J. Forstner, Louise Frisen, Mark A. Frye, Sébastien Gard, Julie S. Garnham, Fernando S. Goes, Maria Grigoroiu-Serbanescu, Andreas J. Fallgatter, Sophia Stegmaier, Thomas Ethofer, Silvia Biere, Kristiyana Petrova, Ceylan Schuster, Kristina Adorjan, Monika Budde, Maria Heilbronner, Janos L. Kalman, Mojtaba Oraki Kohshour, Daniela Reich-Erkelenz, Sabrina K. Schaupp, Eva C. Schulte, Fanny Senner, Thomas Vogl, Ion-George Anghelescu, Volker Arolt, Udo Dannlowski, Detlef E. Dietrich, Christian Figge, Markus Jäger, Fabian U. Lang, Georg Juckel, Carsten Konrad, Jens Reimer, Max Schmauß, Andrea Schmitt, Carsten Spitzer, Martin von Hagen, Jens Wiltfang, Jörg Zimmermann, Till F.M. Andlauer, Andre Fischer, Felix Bermpohl, Philipp Ritter, Silke Matura, Anna Gryaznova, Irina Falkenberg, Cüneyt Yildiz, Tilo Kircher, Julia Schmidt, Marius Koch, Kathrin Gade, Sarah Trost, Ida S. Haussleiter, Martin Lambert, Anja C. Rohenkohl, Vivien Kraft, Paul Grof, Ryota Hashimoto, Joanna Hauser, Stefan Herms, Per Hoffmann, Esther Jiménez, Jean-Pierre Kahn, Layla Kassem, Po-Hsiu Kuo, Tadafumi Kato, John Kelsoe, Sarah Kittel-Schneider, Ewa Ferensztajn-Rochowiak, Barbara König, Ichiro Kusumi, Gonzalo Laje, Mikael Landén, Catharina Lavebratt, Marion Leboyer, Susan G. Leckband, Alfonso Tortorella, Mirko Manchia, Lina Martinsson, Michael J. McCarthy, Susan L. McElroy, Francesc Colom, Vincent Millischer, Marina Mitjans, Francis M. Mondimore, Palmiero Monteleone, Caroline M. Nievergelt, Markus M. Nöthen, Tomas Novák, Claire O’Donovan, Norio Ozaki, Andrea Pfennig, Claudia Pisanu, James B. Potash, Andreas Reif, Eva Reininghaus, Guy A. Rouleau, Janusz K. Rybakowski, Martin Schalling, Peter R. Schofield, Barbara W. Schweizer, Giovanni Severino, Paul D. Shilling, Katzutaka Shimoda, Christian Simhandl, Claire M. Slaney, Alessio Squassina, Thomas Stamm, Pavla Stopkova, Mario Maj, Gustavo Turecki, Eduard Vieta, Julia Veeh, Biju Viswanath, Stephanie H. Witt, Adam Wright, Peter P. Zandi, Philip B. Mitchell, Michael Bauer, Martin Alda, Marcella Rietschel, Francis J. McMahon, Thomas G. Schulze, Bernhard T. Baune, Klaus Oliver Schubert, Azmeraw T. Amare

## Abstract

**Background:** The predictive power of polygenic scores (PGSs) for lithium treatment response in bipolar disorder (BD) remains limited.

**Aim:** To enhance prediction of lithium responsiveness by developing a multi-trait PGS (mt-PGS) combining genetic information from multiple phenotypes implicated in lithium response and/or BD aetiology.

**Methods:** We analysed data collected from BD patients who had received lithium treatment for at least six months and participated in the International Consortium on Lithium Genetics (ConLi^+^Gen, N=2,367) study. The ALDA scale was used to assess lithium responsiveness, and treatment outcome was defined as continuous ALDA score (0-10) and categorical outcome (favourable ≥7 vs unfavourable response). PGSs were calculated for 59 phenotypes grouped into five clinical-biological clusters: clinical lithium exemplar (#22 phenotypes), cardiometabolic (#17), autoimmune/inflammatory (#5), neurocognitive (#8) and renal function (#7). We applied cross-validated machine learning regression approaches in both outcomes within each cluster, and the selected features from each cluster were subsequently combined to construct the final mt-PGS models. Model performance was assessed using explained variance (R²) for the continuous outcome, and McFadden’s pseudo-R² as well as standard classification model parameters for the categorical outcomes.

**Results:** The mt-PGS explained 5.07% (continuous outcome) to 9.02% (categorical outcome) of the interindividual variability in lithium responsiveness. Classification accuracy (AUC) for the categorical outcome was 68.13% (95% CI: 64.86, 71.77). Of the five clusters, the PGSs for clinical lithium exemplar phenotypes were most strongly associated with lithium responsiveness, accounting for 2.97%-6.20% of its variability.

**Conclusions:** By integrating polygenic scores for multiple relevant phenotypes, predictive accuracy for lithium response improved up to nine-fold compared to single-trait methods. Future research incorporating larger, more diverse populations and combining genetic scores with clinical data holds promise for further enhancing prediction and advancing clinical implementation.

## Background

Bipolar disorder (BD) is one of the most severe and prevalent mental illnesses, affecting about 40 to 50 million people worldwide (1), with an estimated 15–20% dying by suicide (2). It is a complex disorder characterized by recurrent hypomanic or manic and depressive episodes and is associated with significant somatic and psychiatric comorbidity (3). Lithium remains the gold standard and first-line medication for patients with BD for over 70 years (4–6), effectively treating acute episodes of illness, preventing relapses, and demonstrating anti-suicidal properties (7–9). However, not all BD patients experience sustained clinical benefits. Studies indicate that only about 30% of patients achieve full clinical remission with lithium monotherapy, while 30-40% exhibit minimal clinical response (10, 11). Clinical causes and predictors of this response heterogeneity have been identified and summarized into a “clinical exemplar” for optimal lithium response (12–14). Yet, these clinical variables alone do not fully explain interindividual variability (15), and in practice are difficult to comprehensively elicit from patients at the point of treatment decision. Therefore, the field has pursued the search for an objective biological predictor for the lithium-responsive phenotype, which could support clinicians and patients in their decision-making.

Previous studies have developed polygenic scores (PGSs) to predict lithium responsiveness in BD, focusing on genetic risk profiles for related psychiatric conditions such as schizophrenia (SCZ) (16), major depressive disorders (MDD) (17), attention-deficit hyperactivity disorder (ADHD) (18) and lithium-responsiveness phenotype itself (19, 20). While these PGSs have shown statistically significant associations with lithium treatment outcomes, each individual PGS explains only a small portion of the variability in lithium response — typically between 0.18% and 2.6%, indicating limited clinical utility when used alone (21–23).

An emerging approach to improve predictive accuracy is integrating PGSs for multiple phenotypes that share genetic architecture with the phenotype of interest (24–27). For example, Schubert et al. meta-analysed the PGSs for MDD and SCZ and found a two-fold increase in predictive capability compared to single-trait PGSs (24). Another study by Albinana and colleagues combined 937 PGSs derived from multiple diseases and conditions that enhanced predictive accuracy for psychiatric diagnoses such as ADHD, affective disorder, autism spectrum disorder, BD, anorexia nervosa and SCZ. Using this methodology, the explained variance for each diagnosis was four to nine-fold larger, compared with a single-disorder PGS (25). Similarly, Krapohl et al. used 81 Genome-wide association studies (GWASs) of cognitive, medical, and anthropometric phenotypes to predict educational achievement, general cognitive ability and body mass index (BMI). They found that the combined PGSs predicted 10.9% variance in educational achievement, 4.8% in general cognitive ability, and 5.4% in BMI – representing increases of 10%, 10%, and 60%, respectively, compared to the best-performing single-score predictions (26). These examples highlight the potential value of harnessing the shared genetic architecture of multiple phenotypes to improve the predictive power of PGSs for complex traits such as lithium response in BD.

## Aims

In the current study, we applied supervised machine learning methods within a multi-trait polygenic score (mt-PGS) framework to predict lithium responsiveness in patients with BD. By integrating PGSs from multiple phenotypes previously implicated in BD pharmacology and/or aetiology, we evaluated whether the mt-PGS approach would improve predictive performance. Additionally, we explored the relative contribution of each phenotype-specific PGS to lithium response, providing insights into its genetic relationship with lithium response phenotype.

## Methods

This study adheres to the Transparent Reporting of a multivariable Prediction model for Individual Prognosis Or Diagnosis – Artificial Intelligence (TRIPOD-AI) guideline (28) to report the findings. The completed TRIPOD-AI checklist is provided in the *supplementary material*.

### Data Source and Patient Population

We utilized data from the International Consortium on Lithium Genetics (ConLi^+^Gen; http://www.conligen.org/), a global initiative investigating the genetic basis of lithium response in BD. For this analysis, we included 2,367 patients of European ancestry who received lithium and were followed up for a minimum of six months. Participants were recruited from Europe, North America, Asia, and Australia between 2008 and 2013 (29). Detailed characteristics of this cohort, including clinical and demographic information, are available elsewhere (11).

### Target outcome measure

The ALDA scale, a validated retrospective criteria of long-term treatment response in research subjects, was used to assess patient’s response to lithium treatment (30, 31). This scale quantifies the degree of improvement during lithium therapy by combining changes in the frequency and severity of mood symptoms into an A score (ranging from 0 to 10). To account for potential confounding factors influencing symptom improvement, the scale incorporates a B score, which considers five confounders, each scored 0, 1, or 2. The total score is then derived by subtracting the total B score from the A score. The detailed procedures for ALDA scale measurement are described elsewhere (29–31). For this study, lithium treatment outcome was defined both as a continuous outcome (total ALDA score) and as a categorical outcome, with patients having a total score of 7 or higher classified as “favourable responders,” and those with a score less than 7 classified as “unfavourable responders” (11, 31).

An overview of the steps involved in developing mt-PGS and the data analysis procedures is outlined in Figure 1.

**Figure 1:**
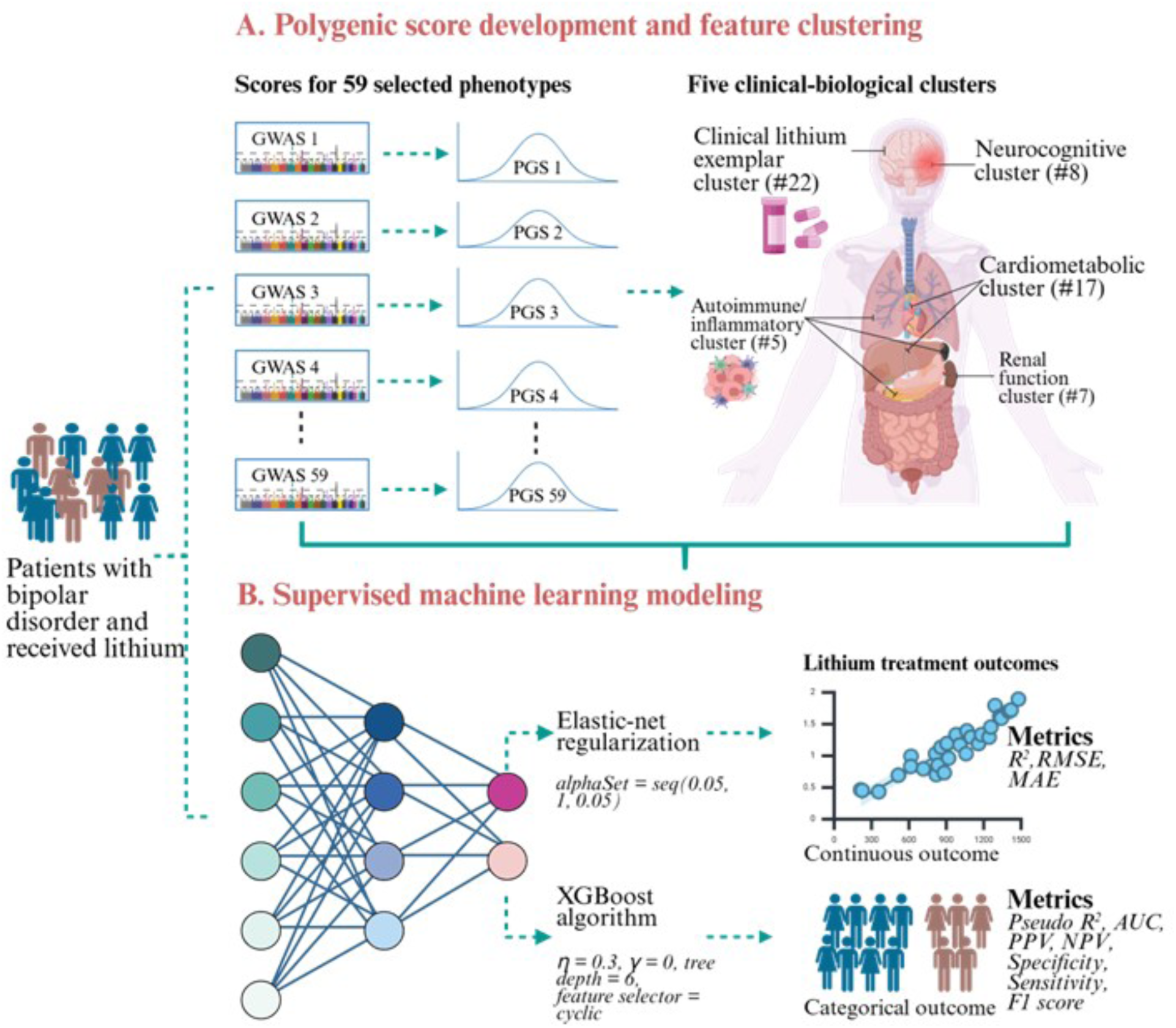
The detailed steps for developing mt-PGS and data analysis procedures. *Abbreviations:* mt-PGS = Multi-trait polygenic score; GWAS = Genome-wide association study; ConLi^+^Gen = The International Consortium on Lithium Genetic; PGS = Polygenic score; R^2^ = Explained variance; RMSE = Root mean squared error; MAE = Mean accuracy error; AUC = Area under the curve; PPV = Positive predictive value; NPV = Negative predictive value

#### Step-1: Selection of phenotypes

For this study, we identified 59 phenotypes that have previously been implicated in and/or potentially share a genetic architecture with pharmacology or aetiology of BD. These included psychiatric disorders and clinical-behavioural phenotypes (32, 33), cardiometabolic phenotypes (34–36), cognitive functions (37), autoimmune and inflammatory conditions (38) and renal function biomarkers (39). A detailed literature review of each phenotype and its association with BD pharmacology or aetiology is provided in the *supplementary literature review*.

#### Step-2: Computing polygenic scores and building a multi-trait PGS library

To develop PGSs, we extracted publicly available summary statistics (discovery data) for the selected phenotypes from the GWAS catalogue [https://www.ebi.ac.uk/gwas/](40), GWAS Atlas [https://atlas.ctglab.nl/] (41), GIANT consortium [https://portals.broadinstitute.org/collaboration/giant/index.php/GIANT_consortium], CNCR - CTG lab [https://cncr.nl/ctg/], and Psychiatric Genomics Consortium [https://www.med.unc.edu/pgc]. The detailed characteristics of the GWAS summary statistics used are provided in the *supplementary table 1*.

Before developing the PGSs, genotype data from the ConLi^+^Gen consortium (target sample) underwent standard quality control (QC) and imputation procedures, as previously described (11, 16, 17, 20, 24, 42). A brief summary of the genotyping, QC, and imputation steps is provided in the *supplementary methods*.

Using the GWAS summary statistics described above, the QC-filtered individuals and SNPs from the ConLi^+^Gen sample and the precomputed linkage disequilibrium (LD) of the 1000 Genome European external reference panel (43), PGSs for each phenotype were computed. The PRS-CS polygenic scoring method was employed to construct the PGS for each phenotype (44). Subsequently, we constructed an agnostic PGSs library for the selected phenotypes. Each score was standardized to a zero mean and unit variance. Detailed information on the computation of PGSs is available in the *supplementary methods*.

#### Step-3: Phenotype clustering for dimensionality reduction

To minimise the overfitting and to ensure precise estimation of key parameters in the prediction models, we determined the optimal number of features in each cluster using the “*pmsampsize*” R package (45), based on our previous machine learning model employed in a similar dataset (46). In that model, an explained variance (R^2^) of 7.1%-9.3% with a standard error of 3.14 was reported in the continuous lithium treatment response model. Using a larger shrinkage parameter (*λ = 0.95*), we found that 10-18 features were optimal for a sample size of 2367 and 660 responders. Based on this guide, we adopted an expertise-driven clustering approach (47) and grouped the PGSs into five clinical-biological clusters: psychiatric disorders and clinical-behavioural phenotypes or (“*clinical lithium exemplar*”, #22); cardiometabolic *(#17)*; *autoimmune/inflammatory* (#5); *neurocognitive (#8)*, and *renal function* (#7) clusters. Full lists of phenotypes in each clinical-biological cluster are provided in the *supplementary table 2*.

#### Step Four: Developing machine learning models

Prior to the mt-PGS modelling, we performed a feature reduction analysis within each cluster using elastic-net regression for the continuous outcome and eXtreme Gradient-Boosted decision tree-based ensembles (XGBoost) for the categorical outcome. Features retained from each cluster were then combined for the final mt-PGS analysis. For both categorical and continuous outcomes, models were trained and evaluated using five-fold nested cross-validation. The dataset was partitioned into outer and inner folds. Inner folds were used to tune hyperparameters and minimize cross-validation error, while the model was trained on the entire inner training set and evaluated on left-out data from the outer fold. This process was repeated across all outer folds, and then the ground truth was compared to pooled, unseen test predictions (48). Detailed modelling procedures for each outcome are described below.

##### A. Continuous outcome model

Elastic-net regularization regression was employed to optimize the hyperparameters and estimate the generalization error in predicting the continuous ALDA score. Elastic-net is a hybrid approach to regularization that blends *L1* and *L2* penalties, allowing coefficient shrinkage for feature selection while effectively addressing multicollinearity (49). We used *n_outer_folds = 5, alphaSet = seq(0.05, 1, 0.05)* hyperparameters for the elastic-net models. The predictive performance of models was assessed using R^2^, root mean squared error (RMSE) and mean absolute error (MAE) for each cluster and the final mt-PGS models. We ranked the contribution of features to the continuous outcome based on their absolute standardized coefficient in the final mt-PGS model.

##### B. Categorical outcome model

The XGBoost algorithm was selected (50) to predict the categorical outcome, as it has demonstrated superior performance in mood disorder classification studies (51). The following hyperparameters were used: *eta = 0.3, gamma = 0, tree depth = 6, feature selector = cyclic.* Model discrimination, the accuracy of classification models, was evaluated using area under the curve (AUC), balanced accuracy, accuracy, negative and positive predictive values, sensitivity, specificity, and F1 score. The *boost R* package was used to bootstrap a 95% confidence interval for prediction performance metrics with 500 bootstrap replicates and the “*perc*” method (52). Class weights were applied based on the ratio of responders and non-responders to achieve balanced predictions for imbalanced data and to assign greater importance to misclassifications of the underrepresented class during optimization. The McFadden pseudo R^2^ was calculated (53) and the reported observed R^2^ was transformed to a liability scale (54), taking into account lithium responsiveness prevalence of 30% (11, 55) and the proportion of lithium responders to non-responders in the target cohort. Finally, the importance of features was ranked using the mean absolute SHapley Additive exPlanation (SHAP) value, which determines the contribution of each feature to the categorical outcome in the final mt-PGS model. The Shapley value is the average marginal contribution of a feature value across all possible coalitions in the XGBoost models (56).

All machine learning analyses were conducted in R (version 4.3.1) (57) using the *Caret* package (58).

### Ethics declarations

The Ethics Committee at the University of Heidelberg provided central ethical approval for the consortium, and written consent was obtained from all participants according to the study protocols of each of the participating sites and their institutions. All procedures were performed according to the Declaration of Helsinki guidelines (11, 38).

## Results

### Cohort characteristics

A total of 2,367 patients with BD who had been treated with lithium for at least six months were included in this study. Nearly 60% of the participants were females; and the mean age was 47.53±13.73 years. Of the total sample, 660 patients (27.9%) responded favourably to lithium treatment, and the mean ALDA total score was 4.12±3.15. Detailed sample characteristics of the cohort are provided in previous publications (11, 16, 17, 20, 24).

### Multi-trait polygenic model for Lithium response (Continuous outcome)

The final mt-PGS model combining PGSs from 25 phenotypes across five clusters explained 5.07% of the variance in lithium responsiveness (RMSE = 3.24, MAE = 2.82; see Table 1 for full details). The top ten contributing PGSs for the final mt-PGS model were scores for lithium responsiveness, BD, major depression, heart failure, COVID-19 susceptibility, asthma, agreeableness, chronic pain, and ADHD (*See Figure 2)*

**Figure 2:**
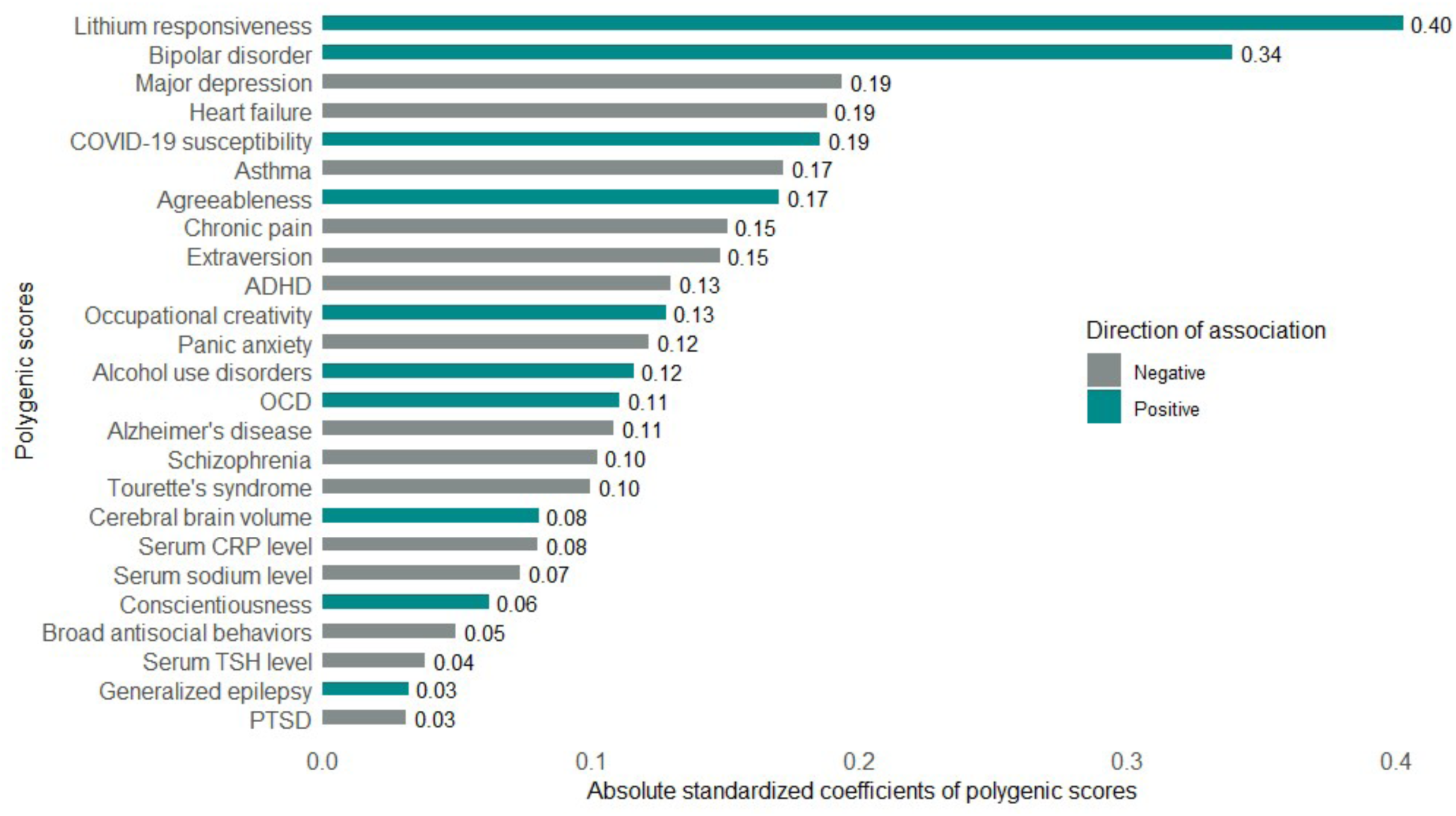
Retained polygenic scores in the final mt-PGS model of continuous outcome. *Abbreviations:* mt-PGS = Multi-trait polygenic score; ADHD = Attention-deficit hyperactivity disorder; OCD = Obsessive compulsive disorder; CRP = C-reactive protein; TSH = Thyroid stimulating hormone; PTSD = Posttraumatic stress disorder

**Table 1:** Continuous outcome model parameters in cluster-wise and combined cluster models.

| Model for | Contributing features | Model parameters |  |  |
| --- | --- | --- | --- | --- |
|  |  | RMSE | MAE | R<sup>2</sup><br>(%) |
| Clinical lithium exemplars | Lithium responsiveness, BD, major depression, schizophrenia, agreeableness, extraversion, conscientiousness, ADHD, panic anxiety, alcohol use disorders, OCD, broad anti-social behaviours, PTSD and Tourette's syndrome | 3.27 | 2.85 | 2.97 |
| Neurocognitive | Chronic pain, occupational creativity, Alzheimer's disease, cerebral brain volume and generalised epilepsy | 3.32 | 2.92 | 0.72 |
| Autoimmune/Inflammatory | Asthma, COVID-19 susceptibility and serum CRP level | 3.30 | 2.89 | 0.71 |
| Cardiometabolic | Heart failure and serum TSH level | 3.31 | 2.91 | 0.58 |
| Renal function | Serum sodium level | 3.33 | 2.92 | ~0.01 |
| Final mt-PGS model | All the above | 3.24 | 2.82 | 5.07 |

*Abbreviations:* BD = Bipolar disorder; ADHD = Attention-deficit hyperactivity disorder; OCD = Obsessive-compulsive disorder; PTSD = Posttraumatic stress disorder; TSH = Thyroid stimulating hormone; RMSE = Root Mean Squared Error; MAE = Mean Accuracy Error; R<sup>2</sup> = Explained variance; mt-PGS = Multi-trait polygenic score

Per-cluster mt-PGS modelling showed that the clinical lithium exemplar (#14 phenotypes), neurocognitive (#5), autoimmune/inflammatory (#3), and cardiometabolic (#2) clusters explain 2.97%, 0.72%, 0.70%, and 0.58% of the variability in lithium responsiveness, respectively. The *renal function cluster* contributed the least to the overall model, with only the PGS for serum sodium level accounting for ∼0.01% of the variance in lithium responsiveness. A detailed breakdown of the variance explained by cluster-wise and the final mt-PGS model is provided in *Table 1*.

### Multi-trait polygenic model for Lithium response (Categorical outcome)

Of the 59 PGSs initially considered, 23 were retained in the final mt-PGS model, collectively explaining 9.02% of the variance in lithium responsiveness on the liability scale (McFadden pseudo-R²; SE = 0.018) and resulted in a prediction accuracy of 68.13% (95% CI: 64.86, 71.77) for lithium responders. The top contributing features in order of importance were: the PGS for lithium responsiveness is the most dominant feature contributor to the final mt-PGS model parameters for the categorical outcome, followed by agreeableness, chronic pain, BD, panic anxiety, ADHD, serum CRP level, Tourette’s syndrome, COVID-19 susceptibility and major depression *(See Figure 3)*.

**Figure 3:**
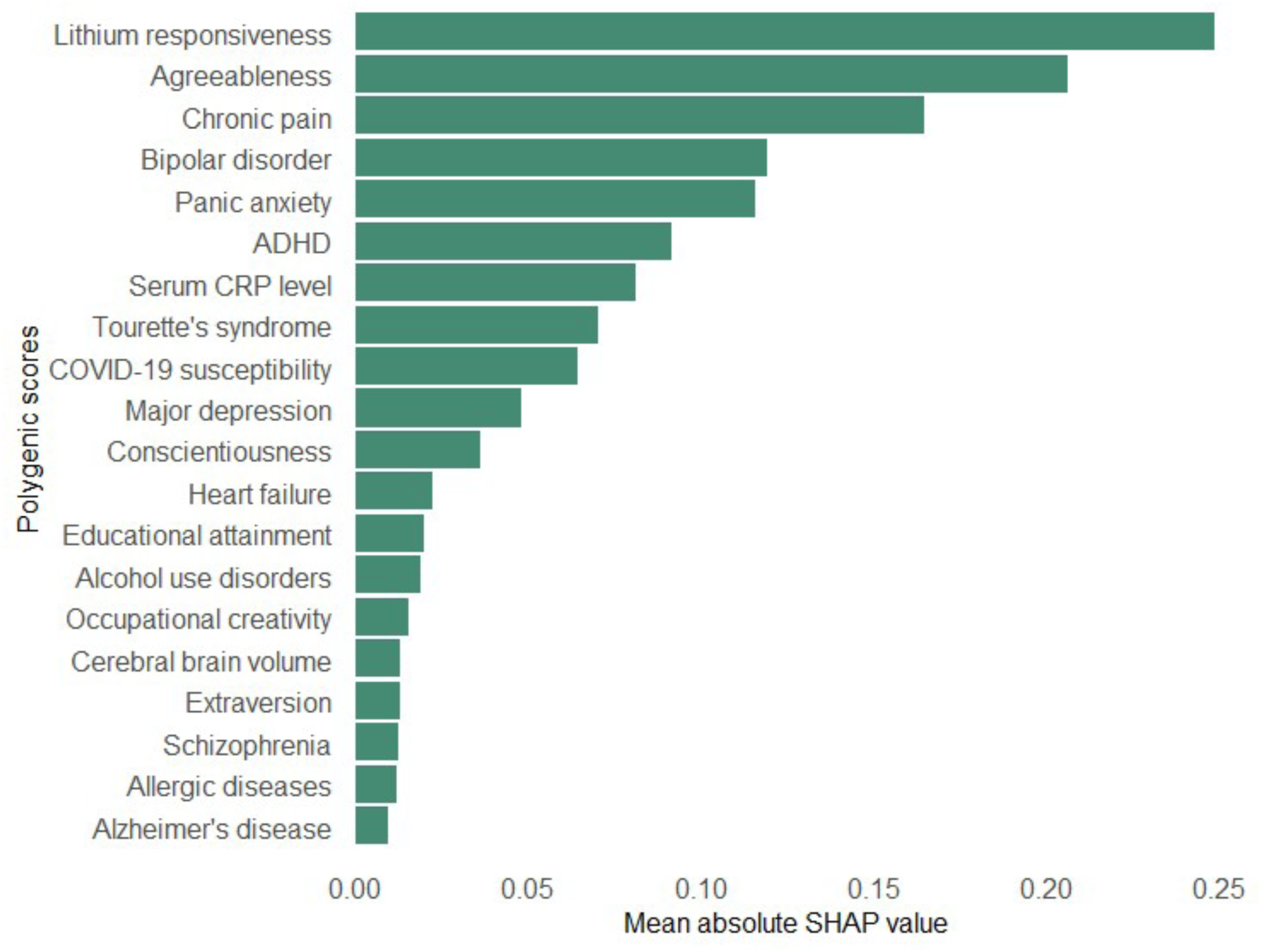
The top 20 polygenic scores contributing to the final mt-PGS model of categorical outcome. *Abbreviations:* mt-PGS = Multi-trait polygenic score; ADHD = Attention-deficit hyperactivity disorder; CRP = C-reactive protein *Legend:* Bar plot of polygenic scores importance, ranked from the most to the least (top to bottom).

Cluster-wise analyses showed that the PGSs from the clinical lithium exemplar, autoimmune/inflammatory, neurocognitive, and cardiometabolic clusters explained 6.20%, 1.88%, 1.64%, and 0.68% of the variance in lithium responsiveness in liability scale, with corresponding AUCs of 62.02% (95% CI: 58.06–65.63), 55.37% (95% CI: 51.62–59.30), 51.99% (95% CI: 48.43–55.98), and 52.43% (95% CI: 48.79–56.21), respectively. No phenotype in the *renal function cluster* contributed meaningfully to the lithium responsiveness (categorical outcome). A full summary of the categorical outcome model parameters in cluster-wise and final mt-PGS models is provided in *Supplementary Table 3*.

## Discussion

This study demonstrates that mt-PGS approach, which integrates genetic risk information from a broad spectrum of phenotypes implicated in lithium pharmacology and the aetiology BD, substantially improves the prediction of lithium responsiveness. Our mt-PGS explained 9.02% of the variance in the categorical and 5.07% in the continuous lithium prediction model, with a maximum AUC of 68.13% (95% CI: 64.86, 71.77). These results represent a substantial improvement (four to nine-fold increase) over single-trait PGS models (e.g., schizophrenia, major depression, and bipolar disorder), which explained only 1–2.6% of the variance in lithium responsiveness (16–18, 20, 24).

The superior performance of the mt-PGS model is likely due to several factors. First, by leveraging polygenic information from multiple, genetically correlated phenotypes, the model captures a broader and more relevant spectrum of genetic variance (13, 59). This approach aligns with recent research showing that multi-trait models, especially those integrating traits with shared biological pathways, consistently outperform single-trait models in complex trait prediction (21, 24, 27, 60, 61). Second, the inclusion of PGSs from medical and behavioural phenotypes that are already known to be associated with psychiatric conditions, as well as pharmacogenomic phenotypes, increases statistical power and biological specificity (62).

Polygenic scores for phenotypes within the clinical lithium exemplar cluster emerged as the most strongly associated features in predicting lithium responsiveness, with R² of 2.97% in the continuous outcome model and McFadden pseudo-R² of 6.02% in the categorical outcome model. This strong association is expected, given that the clinical lithium exemplar phenotype shares a similar genetic architecture with both BD and lithium responsiveness, as established in previous genetic studies (13, 59). Of the individual phenotypes contributing to this cluster, PGSs for BD and lithium responsiveness phenotype were positively associated with actual lithium response, echoing earlier researches (20, 63) and supporting the idea that individuals with a higher genetic predisposition for BD and lithium response are more likely to benefit from lithium treatment. Conversely, PGSs for MDD, SCZ, ADHD, panic anxiety, and broad antisocial behavior were negatively associated with lithium responsiveness. This is consistent with clinical observations that BD patients with a family history of these disorders often respond less favorably to lithium (16–18, 24). Additionally, PGSs for agreeableness and conscientiousness personality traits were positively associated with lithium response, possibly reflecting the influence of these traits on treatment adherence and supportive behaviors (64). Agreeableness, in particular, is linked to increased support-seeking, emotional expression, and positive affect, which may facilitate better clinical outcomes (65). In contrast, extraversion was associated with poorer lithium response, suggesting that different personality structures can have distinct effects on treatment efficacy. Collectively, these findings highlight the complex interplay between genetic risk for psychiatric and behavioral traits and the likelihood of responding to lithium, underscoring the potential value of multi-trait polygenic approaches in guiding personalized treatment strategies for BD.

Autoimmune and inflammatory PGSs explained a modest but notable proportion of lithium responsiveness, accounting for 0.71% to 1.88% of the variance. Within this group, PGSs for asthma and serum CRP levels were negatively associated with lithium response, indicating that individuals with higher genetic risk for these inflammatory conditions are less likely to benefit from lithium treatment. This finding aligns with the established role of inflammation in BD, where elevated CRP and chronic low-grade inflammation are linked to greater symptom severity and shorter durations of effective lithium therapy. Lithium’s recognized immune-modulating properties, including its influence on proinflammatory cytokine regulation (66), may underlie some of its therapeutic effects in mood disorders. The negative association between serum CRP PGS and lithium response supports the hypothesis that lithium’s anti-inflammatory action could be a key mechanism in its efficacy, and that individuals with a genetic predisposition to higher inflammation may experience less benefit Furthermore, immune-related gene sets have been shown to be overrepresented in lithium resistance, with activation of inflammatory and adaptive immune pathways observed in non-responders (38).

Interestingly, the study found a significant positive association between the PGS for COVID-19 susceptibility and lithium response. This is perhaps surprising, but lithium has demonstrated antiviral properties including activity against COVID-19, in preclinical studies (68). One possible explanation is that genetic variants conferring susceptibility to COVID-19 may overlap with immune pathways that also enhance lithium’s therapeutic effects, or that individuals with these variants may have an immune profile that is particularly responsive to lithium’s modulatory actions. However, this association warrants further investigation to clarify the underlying biological mechanisms. Overall, these findings highlight the complex interplay between immune function, inflammation, and lithium efficacy, suggesting that genetic risk for immune-related traits may help predict which patients are most likely to benefit from lithium therapy in BD.

Polygenic scores for neurocognitive phenotypes such as occupational creativity, cerebral brain volume, and general epilepsy were found to be positively associated with lithium responsiveness, while PGS for Alzheimer’s disease and chronic pain showed a negative association. These findings are consistent with a growing body of evidence highlighting lithium’s multifaceted neuroprotective effects (69). Long-term lithium treatment has been shown to not only stabilize mood but also to exert neuroprotective actions, including the reversal of neuropathological changes associated with Alzheimer’s disease and the improvement of memory deficits (70). Neuroimaging studies further support these observations, demonstrating that sustained lithium use is linked to increased hippocampal and amygdala volumes and greater cortical thickness (71, 72)—structural changes that may underlie enhanced divergent thinking, a key component of occupational creativity (73).

The positive association between PGS for epilepsy and lithium responsiveness is also notable, as lithium possesses recognized anticonvulsant properties and is sometimes used adjunctively in epilepsy management (74). Conversely, the negative association with chronic pain PGS may reflect lithium’s involvement in pain modulation pathways; lithium has been shown to inhibit neuromodulators such as substance P and vasoactive intestinal peptide, which are implicated in the perception of pain (75). Moreover, lithium is thought to help restore the balance between excitatory and inhibitory neurotransmission and to promote the synthesis of neuroprotective proteins that are often impaired in chronic pain conditions (75, 76). Collectively, these findings suggest that individuals with a genetic predisposition toward greater neurocognitive capacity or epilepsy may derive more benefit from lithium, while those with genetic risk for neurodegenerative or chronic pain conditions may respond less favorably. This underscores the importance of considering neurocognitive and neurological genetic profiles when predicting lithium response and further supports the integration of neuroimaging and multi-omics data in future precision psychiatry research.

We observed a negative association between PGS for heart failure and serum TSH levels with lithium responsiveness. This suggests that individuals with a higher genetic predisposition to heart failure or elevated TSH—a marker of hypothyroidism—are less likely to respond favorably to lithium treatment. Hypothyroidism is a well-documented side effect of long-term lithium therapy, and a high genetic loading for this condition may reduce the therapeutic benefits of lithium (77, 78). While lithium at therapeutic levels has been reported to exert cardioprotective effects and lower the risk of cardiovascular diseases (79, 80), our findings highlight that genetic risk for cardiometabolic dysregulation may negatively influence lithium response. Cardiometabolic diseases such as heart failure have been associated with an increased risk of renal impairment, which can reduce lithium clearance leading to lithium accumulation and toxicity-thereby altering its therapeutic efficacy (81).

Lithium is primarily eliminated from the body via the renal system (82), and its use is well known to be associated with both acute side effects, such as polyuria, and long-term complications, including chronic renal failure (77, 83, 84). Chronic lithium-induced renal disease is typically characterized by a gradual decline in glomerular filtration rate (eGFR) and creatinine clearance (85). In our analysis, we included the PGSs for renal function traits such as eGFR, serum electrolytes, creatinine, a key biomarker of renal function, and vitamin D, which is activated in the kidneys (86). Despite the clear pharmacokinetic relevance of these renal and electrolyte phenotypes to lithium, their combined contribution to the variability in lithium responsiveness was negligible in our study. This finding suggests that, while renal function is critical for lithium clearance, genetic loading for renal function traits may not be determinant of its therapeutic efficacy.

## Limitations

Our study should be interpreted in light of important limitations. *First*, although the ConLi^+^Gen cohort represents the largest available dataset for studying lithium treatment outcomes in BD, its sample size remains modest compared to cohorts used in other large-scale genetic studies (87). This limitation may reduce the statistical power to generate highly precise PGSs and could affect the stability of our findings (11, 22, 23). *Second*, the application of machine learning analyses to a relatively small sample raises the possibility of overfitting, which can artificially inflate prediction accuracy (48). While we employed nested cross-validation and careful hyperparameter tuning to mitigate this risk, the potential for overestimation of predictive performance cannot be entirely excluded. *Third*, despite our model explaining a significantly greater proportion of the variance in lithium responsiveness compared to previous studies, its predictive accuracy is not yet sufficient for routine clinical application. *Fourth*, our analysis was restricted to individuals of European ancestry, which means the findings may not be generalizable to populations with different genetic backgrounds and could limit the broader applicability of our results. *Fifth*, our PGSs were constructed using GWAS summary statistics, which primarily capture common genetic variants and may miss the influence of rare variants that could also affect lithium response (88). *Sixth*, the PGS for lithium responsiveness was calculated using the ConLi^+^Gen cohort as both the discovery and target dataset, employing a leave-one-country-out procedure. This approach may overestimate the effect of the lithium responsiveness phenotype and therefore requires validation analysis using an independent target cohort. *Finally.* the set of polygenic predictors/phenotypes selected within each cluster may not be exhaustive or fully representative of all features potentially associated with lithium responsiveness, as additional relevant predictors may yet be identified as the field advances. These limitations highlight the need for larger, more diverse cohorts, prospective study designs, and the inclusion of rare variant analyses in future research to further refine and validate predictive models for lithium response in BD.

## Conclusion

In summary, our study highlights that the predictive power of PGSs for lithium responsiveness can be substantially increased by integrating genetic variants from multiple pharmacogenomic and disease-associated traits, rather than relying solely on single-phenotype scores. This multi-trait approach takes advantage of the shared genetic architecture among related phenotypes, thereby capturing a wider and more clinically relevant spectrum of genetic risk factors. The integration of genomics with clinical data has already demonstrated considerable promise; for example, Cearns et al. showed that combining PGSs for MDD and SCZ with clinical variables using machine learning algorithms explained nearly 14% of the variance in lithium responsiveness, outperforming models based on either genetic or clinical data alone (46). Building on this, further improvements in predictive accuracy are likely achievable by adopting mt-PGS approaches in combination with detailed clinical profiles. Looking ahead, future research should prioritize integrating pharmaco-multiomics data—including genomics, metabolomics, neuroimaging, and clinical variables—with the goal of developing even more robust and personalized predictive models. Such integrative and data-driven strategies hold great promise for advancing precision psychiatry (89), ultimately enabling more tailored and effective treatment selection for individuals with BD and other complex psychiatric conditions.

## Data availability/sharing

**All data used in this analysis is available to ConLi^+^Gen members.**

See http://www.conligen.org/ for more information.

## Software and code sharing

Here are the software: **PRS-CS,** see, https://github.com/getian107/PGRScs; for **PLINK 2,** see https://zzz.bwh.harvard.edu/plink/tutorial.shtml. The custom codes used in this study will be made available upon reasonable request to the corresponding author.

## Acknowledgment

The authors are grateful to all patients who participated in the study, and we appreciate the contributions of clinicians, scientists, research assistants, and study staff who helped in the patient recruitment, data collection, and biological sample preparation of the studies. We are also indebted to the members of the ConLi^+^Gen Scientific Advisory Board (http://www.ConLi+Gen.org/) for critical input over the course of the project. The analysis of this study was carried out using the high-performance computational (HPC) capabilities of the University of Adelaide’s Phoenix Supercomputer https://www.adelaide.edu.au/phoenix/.

## Author contributions

AT Amare conceived and designed the project and secured a fellowship to lead the study. NT Sharew developed the research proposal, conducted the statistical analysis, interpreted the findings, and drafted the manuscript. AT Amare, SR Clark, KO Schubert and S Hartmann provided supervision and critically reviewed the data analysis steps and manuscript draft. All authors contributed genetic and clinical data, provided feedback, and made significant intellectual contributions to the manuscript.

