## Supplementary materials for "Using multi-trait polygenic scores to predict lithium responsiveness in patients with bipolar disorder"

### Supplementary methods

#### Genotyping, quality control, and imputation procedures for the ConLi$^+$Gen sample

DNA was extracted from peripheral blood samples collected at 22 participating sites, and samples were genotyped using either Affymetrix or Illumina SNP arrays (1). Prior to imputation, quality control (QC) procedures were implemented on the genotype data using PLINK version 1.9 (2). SNPs with a poor genotyping rate (< 95%), strand ambiguity (A/T and C/G SNPs), a minor allele frequency (MAF) less than 1%, and SNPs deviated from Hardy-Weinberg Equilibrium (HWE) ($p < 10^{-6}$) were removed. Individuals with sex inconsistencies between the documented and genotype-derived sex and genetically related were also excluded. The genotypic and quality control details of the ConLi$^+$Gen cohort are available elsewhere (1).

The genotype data passing QC were imputed in the Michigan server separately for each genotyping platform using the Haplotype Reference Consortium (HRC) reference panel comprising broadly European haplotypes at 39,235,157 SNPs (3). For each cohort, imputation quality procedures were implemented and excluded SNPs of low-frequency (MAF) < 1%) and low-quality (imputation quality score $R^2 < 0.6$). Then, genotype calls for the filtered SNPs were derived and merged using PLINK from the imputed dosage score (2).

#### Polygenic score computation procedures

The polygenic risk score with continuous shrinkage prior (PRS-CS) polygenic scoring method was employed to construct the polygenic score (PGS) for each phenotype (4) using GWAS summary statistics described below and the precomputed linkage disequilibrium (LD) of the 1000 Genomes European external reference panel(5). PRS-CS employs a Bayesian regression framework, incorporating continuous shrinkage priors on effect sizes and accounting for linkage disequilibrium (LD) among SNPs(4). Continuous shrinkage applies a proportionate reduction to the coefficient estimates of SNPs based on their strength of association. This process yields a pruned set of SNPs with their posterior effect sizes for downstream analysis (4). Genetic data from 1000 Genomes Project Phase 3 was used as an external panel of LD pattern and global shrinkage parameters *(--phi=1e-2)* with other default options to estimate a single PGS for each phenotype. The PGS for each phenotype was then computed, summing up the dosages of effect alleles (ranging from 0 to 2) for independent genotyped SNP multiplied by corresponding posterior SNP effect sizes across the genome in each individual. The PGS was then computed in PLINK version 2(6).

### Supplementary literature review

#### Literature review on the involvement of selected phenotypes in BD pharmacology and aetiology

Several studies have reported shared genetic architecture between bipolar disorder (BD) and other phenotypes, including psychiatric disorders and clinical-behavioural phenotypes (7, 8), cardiometabolic phenotypes (9-11), neurocognitive functions (12), autoimmune and inflammatory conditions (13) and renal function biomarkers (14). Here, we discuss the involvement of these phenotypes and their shared genetic architecture with BD and lithium response, grouped in five clinical-biological clusters.

*Cluster One: Clinical lithium exemplar phenotypes*

Clinical lithium exemplar phenotypes refer to those clinical phenotypes identified through clinical interviews that reliably predict lithium responsiveness based on their characteristics (15, 16). The profiles of clinical exemplars were distinct and consistent with previous phenotypic research on lithium responders. The genetic separability of clinical lithium exemplars associated with lithium response and shared genetics architecture with BD demonstrate the biological validity of the detailed clinical evaluation, whose predictive utility has been previously established (17, 18). This cluster encompasses BD, Major depressive disorder (MDD), schizophrenia (SCZ), alcohol use, obsessive-compulsive disorder, post-traumatic stress disorder, panic anxiety, broad antisocial behavior, anorexia nervosa, attention-deficit hyperactivity disorder (ADHD), autism spectrum disorder, Tourette's syndrome, treatment-resistant schizophrenia, coffee consumption, personality traits, insomnia, as well as responses to psychotropic medications, including lithium responsiveness and responses to antidepressants (17, 19-23).

*Cluster Two: Cardiometabolic phenotypes*

Cardiometabolic phenotypes such as low total cholesterol, low high-density lipoprotein, high triglycerides, high glycated hemoglobin, low systolic blood pressure, high BMI, stroke, coronary artery disease, and type 2 diabetes are significantly associated with an increased risk of bipolar disorder (24). Apolipoprotein A1 has been reported to have a central role in lipid metabolism by mediating cholesterol transport and inflammatory, immunologic and vasodilatory pathways (25, 26), and an increased apolipoprotein A1 has been a stronger prognostic marker for cardiovascular diseases (27). The shared genetic architecture between cardiometabolic phenotypes and mood disorders has been reported in several studies (10, 11).

For example, BD shares pleiotropic genes—such as *WBP1L*, *SERBP1P3*, and *HDAC9*—with coronary artery disease, and with type 2 diabetes it shares *PCBD2*, *SFMBT1*, and *ABCB9*, along with pathways involving chromatin-modifying enzymes, alpha-linolenic acid, linoleic acid metabolism, antigen processing, and presentation with cardiometabolic phenotypes (28). Accumulated evidence suggested that thyroid dysfunction is associated with BD (29, 30) and lithium has been associated with alterations in the thyroid gland, contributing to the development of hypothyroidism and goiter through thyroid-releasing hormone (31).
Given the association between cardiometabolic and BD, cardiometabolic disorders such as heart failure, aortic stenosis, heart rate variability, hypertension, metabolic syndrome, type 2 diabetes, systolic blood pressure, and hypothyroidism were assigned to this cluster. Additionally, cardiometabolic biomarkers - including thyroid-stimulating hormone, high-density lipoprotein, low-density lipoprotein, serum glucose level, serum total cholesterol level, serum triglyceride level, HbA1C, Apolipoprotein A1 and BMI – were included.

*Cluster Three: Autoimmune and inflammatory phenotypes*

Bipolar disorder has been associated with immune dysfunction, and lithium shows antiviral and immune cell regulatory properties on top of its mood-stabilizing effects (32). Studies have also revealed a significant genetic correlation between BD and immune-related disease, and variations in multiple inflammatory and adaptive immune processes might modestly contribute to the response to lithium treatment in patients with BD (33). Immune-related genes such as *CNTNAP5, DSP*, *NFIB*, *BMPR1A*, and *HAS3* showed pleiotropy for BD, and polygenic scores for immune-related phenotypes were associated with lithium response (13). Inflammatory biomarkers, including interleukins, interferon ϒ, tumor necrosis factor (TNF), and chemokines are elevated, and anti-inflammatory biomarkers are reduced in patients with BD (34), and lithium can impact inflammation (35). Thus, immune and inflammatory phenotypes such as allergic diseases, autoimmune disorders, asthma, COVID-19 and C-reactive protein level phenotypes were grouped in this cluster.

*Cluster Four: Neurocognitive phenotypes*

Neurocognitive impairments are common and can predict functional outcomes in patients with BD (36, 37). These impairments cut across multiple domains such as memory, attention, verbal learning, executive functions, and social cognition in these patients (38). Brain imaging studies reported an abnormal increase of cortical thickening in the medial occipital cortex and central sulcus. The medial occipital cortex and central sulcus form a bimodal visual network (39) in which the neuroanatomy and function of the visual network, perception and emotion recognition (40), visual processing (41) and cognitive process (42) have been implicated in BD, and alterations in the identifications and evaluation of emotion are the hallmarks of BD (43). These studies denoted that brain neural networks involved in sensory perception and emotion processing may play a role in the progressive features of BD. Genetics studies have also reported a negative association between the PGS for BD and activation in the visual association context during facial affect processing, and in contrast, it was associated with failure to deactivate the ventromedial prefrontal regions of the default mode network during working memory processing (44). Chronic pain has been implicated in neurocognitive dysfunction through genetic and neurobiological pathways, with GWAS-based pathway and gene-level analyses identifying neurogenesis and synaptic plasticity as key mechanisms (45). The shared genetic correlations between chronic pain and reduced regional grey matter morphology further support its relevance to BD-related neurocognitive impairment (46). Following this, Alzheimer's disease, educational attainment, intelligence, occupational creativity, cerebral brain volume, hippocampal volume, chronic pain, and generalized epilepsy were assigned to this cluster.

*Cluster Five: Renal function phenotypes*

Lithium is primarily excreted by kidneys, and studies have shown that lithium is associated with a modest decline in renal function and an increased risk of end-stage renal failure (47, 48). A retrospective study by Close and colleagues found that bipolar patients taking lithium were 2.5 times more likely to develop renal failure compared with lithium non-users (49). Similarly, a case-control study reported that lithium users had 6.9% higher plasma creatine concentration and 7.3% lower estimated glomerular filtration rate (eGFR) compared with patients with other psychiatric conditions (50). A Mendelian randomization study reported a negative causal association between vitamin D and renal function as defined by eGFR (51). Moreover, the higher polygenic score for eGFR has been associated with a higher lithium clearance as measured with eGFR (52). The prevalence of hypercalcemia is higher in patients with renal failure associated, likely due to lithium's inhibitory effect on renal calcium and phosphate secretion (53). Lithium-induced renal dysfunction has also been reported to lead to sodium and potassium depletion, with sodium chloride intake helping to stabilize serum lithium levels in patients with BD (54). So, we have considered renal function biomarkers — including eGFR, sodium, potassium, phosphate, creatinine, calcium, and vitamin D levels in this cluster.

**Supplementary Table 1: Characteristics of the GWAS summary statistics of selected phenotypes**

| Phenotypes | Ancestry | Sample size | Reported loci | PMID | References |
|---|---|---|---|---|---|
| **Major depression** | Multi-ancestry (Majorly European; UKBB and PGC cohorts were considered)) | 500,199 | 102 | 30718901 | Howard et al.,(55) |
| **Bipolar disorder** | European | 413,466 | 64 | 34002096 | Mullins et al.,(56) |
| **ADHD** | European | 225,534 | 27 | 36702997 | Demontis et al.,(57) |
| **Panic anxiety** | European | 10,240 | None | 31712720 | Forstner et al.,(58) |
| **Eating disorders** | European | 72,517 | 8 | 31308545 | Watson et al.,(59) |
| **Posttraumatic stress disorder** | European | 174,659 | 6 | 31594949 | Nievergelt et al.,(60) |
| **Autism spectrum disorder** | European | 46,350 | 5 | 30804558 | Grove et al.,(61) |
| **Tourette's syndrome** | European | 14,307 | 1 | 30818990 | Yu et al.,(62) |
| **Obsessive-compulsive disorder** | European | 9,725 | None | 28761083 | IOCDF-GC and OCGAS), (2018)(63) |
| **Alcohol use disorders** | European | 121,604 | 10 | 30336701 | Sanchez-Roige et al.,(64) |
| **Insomnia** | European (only UKBB cohort) | 386,988 | | 35835914 | Watanabe et al.,(65) |
| **Intelligence** | European | 269,867 | 205 | 29942086 | Savage et al.,(66) |
| **Cerebral brain volume** | European | 27,486 | 85 | 35842455 | Tissink et al.,(67) |
| **Antidepressants response** | European | 5,218 | None | 35712048 | Pain et al.,(68) |
| **Treatment resistance schizophrenia** | European | 85,490 | NR | 35019943 | Pardiñas et al.,(69) |
| **Lithium responsiveness** | European | 2,367 | 1 | 37433967 | Amare et al.,(22) |
| **Broad antisocial behaviour** | European | 93,417 | 1 | 36284158 | Tielbeek et al.,(70) |
| **Schizophrenia** | European (majority) and East Asian | 150,064 | 108 | 25056061 | Schizophrenia Working Group of the PGC, (2014) (71) |
| **Occupational creativity** | European | 241,736 | 25 | 38335777 | Kim et al.,(72) |

| | | | | | |
|---|---|---|---|---|---|
| **Neuroticism** | European | 329,821 | 166 | 29255261 | Luciano et al.,(73) |
| **Agreeableness** | European | 20,669 | 0 | 21173776 | de Moor et al.,(74) |
| **Openness** | European | 20,669 | 1 | 21173776 | de Moor et al.,(74) |
| **Conscientiousness** | European | 20,669 | 1 | 21173776 | de Moor et al.,(74) |
| **Extraversion** | European | 20,669 | 0 | 21173776 | de Moor et al.,(74) |
| **Body mass index** | European | 1,100,000 | 906 | 36581621 | Huang et al.,(75) |
| **Serum TSH level** | European | 247,107 | 158 | 37872160 | Williams et al.,(76) |
| **Type 2 diabetes** | European and Asian | 659,316 | 143 | 30054458 | Xue et al.,(77) |
| **Allergic disease** | European | 102,453 | 33 | 29785011 | Zhu et al.,(78) |
| **Asthma** | European | 90,853 | 6 | 29785011 | Zhu et al.,(78) |
| **Autoimmune diseases** | European | 352,553 | 0 | 34278373 | Glanville et al.,(79) |
| **C-reactive protein** | European | 1,002,898 | 266 | 35459240 | Saredo et al.,(80) |
| **Alzheimer's disease** | European | 788,989 | 75 | 35379992 | Bellenguez et al.,(81) |
| **Educational attainment** | European | 3,037,499 | 3952 | 35361970 | Okbay et al.,(82) |
| **Chronic pain** | European | 380,000 | 76 | 31194737 | Johnston et al.,(83) |
| **Heart failure** | Multiple ancestries (Majorly European) | 1,665,481 | 44 | 36376295 | Levin et al.,(84) |
| **Metabolic syndrome** | European | 291,107 | 93 | 31589552 | Lind (85) |
| **Aortic stenosis and aortic valve calcification** | European | 956,682 | 32 | 38494474 | Thériault et al.,(86) |
| **Systolic blood pressure** | European | 349,328 | 51 | 38459180 | Yang et al.,(87) |
| **Hypertension** | European | 87,981 | 0 | 38459180 | Yang et al.,(87) |
| **Heart rate variability measurement** | European | 46,075 | 13 | 37803156 | Tegegne et al.,(88) |
| **Total cholesterol level** | African American or Afro-Caribbean, Sub-Saharan African | 125,000 | 91 | 37669986 | Kamiza et al.,(89) |

| | | | | | |
|---|---|---|---|---|---|
| **Generalised epilepsy** | European, African unspecified, Asian unspecified | 59,945 | 36 | 37653029 | International League Against Epilepsy Consortium on Complex Epilepsies, (2023)(90) |
| **Hippocampal volume** | European & East Asian | 38,977 | 20 | 37337106 | Liu et al.,(91) |
| **Hypothyroidism** | European | 494,577 | 69 | 36093044 | Mathieu et al.,(92) |
| **ApoA1** | European and few African and south Asian | 323,833 | 454 | 33462484 | Sinnott-Armstrong et al.,(93) |
| **Serum HbA1c level** | European and few African and south Asian | 367,471 | 470 | 33462484 | Sinnott-Armstrong et al.,(94) |
| **Serum potassium level** | European and few African and south Asian | 345,182 | 1 | 33462484 | Sinnott-Armstrong et al.,(94) |
| **Serum phosphate levels** | European and few African and south Asian | 325,141 | 188 | 33462484 | Sinnott-Armstrong et al.,(94) |
| **Serum sodium level** | European and few African and south Asian | 345,171 | 15 | 33462484 | Sinnott-Armstrong et al.,(94) |
| **Serum creatinine level** | European and few African and south Asian | 355,731 | 359 | 33462484 | Sinnott-Armstrong et al.,(94) |
| **Serum triglyceride level** | European and few African and south Asian | 355,577 | 424 | 33462484 | Sinnott-Armstrong et al.,(94) |
| **Serum LDL level** | European and few African and south Asian | 355,197 | 563 | 33462484 | Sinnott-Armstrong et al.,(94) |
| **Serum HDL levels** | European and few African and south Asian | 325,584 | 529 | 33462484 | Sinnott-Armstrong et al.,(94) |
| **Serum calcium level** | European and few African and south Asian | 325,659 | 208 | 33462484 | Sinnott-Armstrong et al.,(94) |
| **eGFR** | European and few African and south Asian | 355,731 | 349 | 33462484 | Sinnott-Armstrong et al.,(94) |
| **Serum glucose level** | European and few African and south Asian | 325,386 | 121 | 33462484 | Sinnott-Armstrong et al.,(94) |
| **Serum vitamin D levels** | European and few African and south Asian | 339,705 | 142 | 33462484 | Sinnott-Armstrong et al.,(94) |
| **COVID-19 susceptibility** | European | 1,348,701 | 0 | 32404885 | COVID-19 Host Genetics Initiative, (2020)(95) |
| **Coffee consumption measurement** | European | 448,204 | 23 | 32193382 | Cole et al.,(96) |

*Abbreviations:* ADHD = Attention deficit hyperactivity disorder; PGC = Psychiatric Genomics Consortium; UKBB = United Kingdom Biobank; IOCDF-GC = International obsessive compulsive disorder (OCD) foundation genetics collaborative; OCGAS = OCD collaborative genetics Association studies; ApoA1 = apolipoprotein ACRP = C-reactive protein; eGFR = estimated glomerular filtration rate; HDL = High-density lipoprotein, LDL = Low density lipoprotein; TSH = Thyroid stimulating hormone

**Supplementary Table 2: Clinical-biological clustering of polygenic scores**

| S.no | Clinical lithium exemplar cluster | Cardiometabolic cluster | Autoimmune & inflammatory cluster | Neurocognitive cluster | Renal function cluster |
|---|---|---|---|---|---|
| **1.** | Bipolar disorder | Heart failure | Allergic diseases | Alzheimer's disease | Serum sodium |
| **2.** | Schizophrenia | Heart rate variability | Autoimmune disorders | Educational attainment | Serum potassium |
| **3.** | Major depression | Hypertension | Asthma | Intelligence | Serum creatinine |
| **4.** | Broad antisocial behaviour | Aortic stenosis | COVID-19 | Occupational creativity | Serum eGFR |
| **5.** | Autism spectrum disorder | Metabolic syndrome | Serum CRP | Cerebral brain volume | Serum calcium |
| **6.** | ADHD | Body mass index | | Hippocampal volume | Serum vitamin D |
| **7.** | Obsessive-compulsive disorder | Type 2 diabetes | | General epilepsy | Serum phosphate |
| **8.** | Post-traumatic stress disorder | Hypothyroidism | | Chronic Pain | |
| **9.** | Tourette's syndrome | Systolic blood pressure | | | |
| **10.** | Panic anxiety | Serum TSH level | | | |
| **11.** | Insomnia | Serum glucose level | | | |
| **12.** | Anorexia nervosa | Serum HDL level | | | |
| **13.** | Alcohol use disorders | Serum LDL level | | | |
| **14.** | Coffee consumption | Serum total cholesterol | | | |
| **15.** | Personality traits: Neuroticism, Openness, Agreeableness, Conscientiousness, Extraversion | Serum triglyceride | | | |
| **16.** | Lithium responsiveness | ApoA1 | | | |
| **17.** | Treatment-resistant schizophrenia | Serum HbA1c | | | |
| **18.** | Non-response to antidepressants | | | | |

*Abbreviations:* ADHD = Attention deficit hyperactivity disorder; HDL = High-density lipoprotein, LDL = Low density lipoprotein; ApoA1 = apolipoprotein A; CRP = C-reactive protein; eGFR = estimated glomerular filtration rate; TSH = Thyroid stimulating hormone

### Supplementary Table 3: Full summary of the categorical outcome model parameters in cluster-wise and final mt-PGS models

| Model for | Contributing features | McFadden pseudo $R^2$(%) | | Classification prediction model parameters | | | | | | | |
|---|---|---|---|---|---|---|---|---|---|---|---|
| | | Observed | Liability scale (SE) | AUC (95%CI) | Accuracy (95%CI) | Balanced accuracy (95%CI) | PPV (95%CI) | NPV (95%CI) | Specificity (95%CI) | Sensitivity (95%CI) | F1 score (95%CI) |
| **Clinical lithium exemplar** | Lithium responsiveness, BD, major depression, schizophrenia, ADHD, alcohol use disorder, panic anxiety, agreeableness, conscientiousness, extraversion, broad anti-social behaviours and Tourette's syndrome | 4.03 | 6.20(0.014) | 62.02 (58.06, 65.63) | 56.20 (53.43, 59.11) | 56.46 (53.47, 59.70) | 39.49 (35.17, 44.06) | 75.22 (71.72, 78.96) | 57.74 (54.06, 61.25) | 59.18 (54.25, 64.77) | 47.34 (43.05, 51.85) |
| **Cardiometabolic** | Heart failure | 1.22 | 1.88 (0.011) | 52.43 (48.79, 56.21) | 50.13 (47.05, 52.93) | 50.30 (47.34, 53.63) | 34.92 (30.78, 38.98) | 71.35 (67.64, 74.97) | 52.28 (48.68, 55.80) | 54.94 (49.28, 60.38) | 42.68 (38.39, 46.68) |
| **Autoimmune/ Inflammatory** | Asthma, allergic diseases, COVID-19 susceptibility and serum CRP levels | 1.07 | 1.64 (0.001) | 55.37 (51.62, 59.30) | 53.28 (50.55, 56.38) | 53.22 (50.18, 56.57) | 38.42 (33.83, 42.77) | 74.15 (70.92, 77.73) | 56.38 (52.89, 59.96) | 56.07 (50.38, 561.52) | 45.89 (41.23, 50.45) |
| **Neurocognitive** | Chronic pain, educational attainment, cerebral brain volume, general epilepsy, Alzheimer's disease and occupational creativity | 0.44 | 0.68 (0.003) | 51.99 (48.43, 55.98) | 51.72 (48.74, 54.86) | 51.03 (48.06, 54.34) | 34.59 (30.27, 38.56) | 70.69 (67.17, 74.98) | 54.91 (51.03, 58.83) | 51.16 (46.01, 56.56) | 41.25 (36.99, 45.26) |
| **Renal function** | NA | 0.02 | 0.03 (0.002) | 48.76 (45.01, 52.37) | 46.90 (43.11, 49.53) | 49.28 (46.23, 52.64) | 30.10 (25.78, 34.28) | 64.20 (60.30, 71.33) | 51.38 (47.73, 55.46) | 53.73 (48.00, 58.91) | 41.62 (37.05, 45.92) |
| **Final mt-PGS model** | All the above | 5.86 | 9.02 (0.018) | 68.13 (64.86, 71.77) | 64.00 (60.49, 66.87) | 63.67 (60.49, 67.06) | 45.08 (40.55 49.95) | 79.38 (76.02, 82.72) | 64.60 (61.65, 67.94) | 62.73 (58.56, 67.78) | 53.37 (48.93, 57.93) |

*Abbreviations:* BD = Bipolar disorder; ADHD = Attention-deficit hyperactivity disorder; CRP = C-reactive protein; $R^2$ = Explained variance; SE = Standard error; AUC = Area under the curve; PPV = Positive predictive value; NPV = Negative predictive value; CI = Confidence interval; mt-PGS = Multi-trait polygenic score

*Legend:* Final mt-PGS model = Combined retained features across each cluster.

### Supplementary completed checklist of TRIPOD-AI guideline

| Section/Topic / evaluation[1] | Item | Development | Checklist item | Reported on page |
|---|---|---|---|---|
| **TITLE** | | | | |
| *Title* | 1 | D;E | Identify the study as developing or evaluating the performance of a multivariable prediction model, the target population, and the outcome to be predicted | 1 |
| **ABSTRACT** | | | | |
| *Abstract* | 2 | D;E | See TRIPOD+AI for Abstracts checklist | 9 |
| **INTRODUCTION** | | | | |
| *Background* | 3a | D;E | Explain the healthcare context (including whether diagnostic or prognostic) and rationale for developing or evaluating the prediction model, including references to existing models | 10-11 |
| | 3b | D;E | Describe the target population and the intended purpose of the prediction model in the context of the care pathway, including its intended users (e.g., healthcare professionals, patients, public) | 10-11 |
| | 3c | D;E | Describe any known health inequalities between sociodemographic groups | 10-11 |
| *Objectives* | 4 | D;E | Specify the study objectives, including whether the study describes the development or validation of a prediction model (or both) | 11 |
| **METHODS** | | | | |
| *Data* | 5a | D;E | Describe the sources of data separately for the development and evaluation datasets (e.g., randomised trial, cohort, routine care or registry data), the rationale for using these data, and representativeness of the data | 12 |
| | 5b | D;E | Specify the dates of the collected participant data, including start and end of participant accrual; and, if applicable, end of follow-up | 12 |
| *Participants* | 6a | D;E | Specify key elements of the study setting (e.g., primary care, secondary care, general population) including the number and location of centres | 12 |
| | 6b | D;E | Describe the eligibility criteria for study participants | 12 |
| | 6c | D;E | Give details of any treatments received, and how they were handled during model development or evaluation, if relevant | 12 |
| *Data preparation* | 7 | D;E | Describe any data pre-processing and quality checking, including whether this was similar across relevant sociodemographic groups | 13 & supp |
| *Outcome* | 8a | D;E | Clearly define the outcome that is being predicted and the time horizon, including how and when assessed, the rationale for choosing this outcome, and whether the method of outcome assessment is consistent across sociodemographic groups | 12 |
| | 8b | D;E | If outcome assessment requires subjective interpretation, describe the qualifications and demographic characteristics of the outcome assessors | NA |
| | 8c | D;E | Report any actions to blind assessment of the outcome to be predicted | NA |
| *Predictors* | 9a | D | Describe the choice of initial predictors (e.g., literature, previous models, all available predictors) and any pre-selection of predictors before model building | 13, 14 & supp |
| | 9b | D;E | Clearly define all predictors, including how and when they were measured (and any actions to blind assessment of predictors for the outcome and other predictors) | 13,14 & supp |
| | 9c | D;E | If predictor measurement requires subjective interpretation, describe the qualifications and demographic characteristics of the predictor assessors | NA |
| *Sample size* | 10 | D;E | Explain how the study size was arrived at (separately for development and evaluation), and justify that the study size was sufficient to answer the research question. Include details of any sample size calculation | 14 |
| *Missing data* | 11 | D;E | Describe how missing data were handled. Provide reasons for omitting any data | 14 |
| *Analytical methods* | 12a | D | Describe how the data were used (e.g., for development and evaluation of model performance) in the analysis, including whether the data were partitioned, considering any sample size requirements | 14-15 |
| | 12b | D | Depending on the type of model, describe how predictors were handled in the analyses (functional form, rescaling, transformation, or any standardisation). | 14-15 |
| | 12c | D | Specify the type of model, rationale[2], all model-building steps, including any hyperparameter tuning, and method for internal validation | 14-15 |

| | | | | |
|---|---|---|---|---|
| | 12d | D;E | Describe if and how any heterogeneity in estimates of model parameter values and model performance was handled and quantified across clusters (e.g., hospitals, countries). See TRIPOD-Cluster for additional considerations[3] | NA |
| | 12e | D;E | Specify all measures and plots used (and their rationale) to evaluate model performance (e.g., discrimination, calibration, clinical utility) and, if relevant, to compare multiple models | 14-15 |
| | 12f | E | Describe any model updating (e.g., recalibration) arising from the model evaluation, either overall or for particular sociodemographic groups or settings | 14-15 |
| | 12g | E | For model evaluation, describe how the model predictions were calculated (e.g., formula, code, object, application programming interface) | 14-15 |
| *Class imbalance* | 13 | D;E | If class imbalance methods were used, state why and how this was done, and any subsequent methods to recalibrate the model or the model predictions | 15 |
| *Fairness* | 14 | D;E | Describe any approaches that were used to address model fairness and their rationale | 15 |
| *Model output* | 15 | D | Specify the output of the prediction model (e.g., probabilities, classification). Provide details and rationale for any classification and how the thresholds were identified | 14-15 |
| *Training versus evaluation* | 16 | D;E | Identify any differences between the development and evaluation data in healthcare setting, eligibility criteria, outcome, and predictors | 14-15 |
| *Ethical approval* | 17 | D;E | Name the institutional research board or ethics committee that approved the study and describe the participant-informed consent or the ethics committee waiver of informed consent | 15 |
| **OPEN SCIENCE** | | | | |
| *Funding* | 18a | D;E | Give the source of funding and the role of the funders for the present study | 28-30 |
| *Conflicts of interest* | 18b | D;E | Declare any conflicts of interest and financial disclosures for all authors | 30-31 |
| *Protocol* | 18c | D;E | Indicate where the study protocol can be accessed or state that a protocol was not prepared | NA |
| *Registration* | 18d | D;E | Provide registration information for the study, including register name and registration number, or state that the study was not registered | NA |
| *Data sharing* | 18e | D;E | Provide details of the availability of the study data | 28 |
| *Code sharing* | 18f | D;E | Provide details of the availability of the analytical code[4] | 28 |
| **PATIENT & PUBLIC INVOLVEMENT** | | | | |
| *Patient & Public Involvement* | 19 | D;E | Provide details of any patient and public involvement during the design, conduct, reporting, interpretation, or dissemination of the study or state no involvement. | NA |
| **RESULTS** | | | | |
| *Participants* | 20a | D;E | Describe the flow of participants through the study, including the number of participants with and without the outcome and, if applicable, a summary of the follow-up time. A diagram may be helpful. | 16 |
| | 20b | D;E | Report the characteristics overall and, where applicable, for each data source or setting, including the key dates, key predictors (including demographics), treatments received, sample size, number of outcome events, follow-up time, and amount of missing data. A table may be helpful. Report any differences across key demographic groups. | 16 |
| | 20c | E | For model evaluation, show a comparison with the development data of the distribution of important predictors (demographics, predictors, and outcome). | 16-17, fig 2 and 3 & tables |
| *Model development* | 21 | D;E | Specify the number of participants and outcome events in each analysis (e.g., for model development, hyperparameter tuning, model evaluation) | 17 |
| *Model specification* | 22 | D | Provide details of the full prediction model (e.g., formula, code, object, application programming interface) to allow predictions in new individuals and to enable third-party evaluation and implementation, including any restrictions to access or re-use (e.g., freely available, proprietary)[5] | 16-17, fig 2 and 3 & tables |
| *Model performance* | 23a | D;E | Report model performance estimates with confidence intervals, including for any key subgroups (e.g., sociodemographic). Consider plots to aid presentation. | 16-17, fig 2 and 3 & tables |
| | 23b | D;E | If examined, report results of any heterogeneity in model performance across clusters. See TRIPOD Cluster for additional details[3]. | NA |
| *Model updating* | 24 | E | Report the results from any model updating, including the updated model and subsequent performance | NA |
| **DISCUSSION** | | | | |

| | | | | |
|---|---|---|---|---|
| *Interpretation* | 25 | D;E | Give an overall interpretation of the main results, including issues of fairness in the context of the objectives and previous studies | 18-22 |
| *Limitations* | 26 | D;E | Discuss any limitations of the study (such as a non-representative sample, sample size, overfitting, missing data) and their effects on any biases, statistical uncertainty, and generalizability | 21-22 |
| *Usability of the model in the context of current care* | 27a | D | Describe how poor quality or unavailable input data (e.g., predictor values) should be assessed and handled when implementing the prediction model | 21-22 |
| | 27b | D | Specify whether users will be required to interact in the handling of the input data or use of the model, and what level of expertise is required of users | 21-22 |
| | 27c | D;E | Discuss any next steps for future research, with a specific view to applicability and generalizability of the model | 21-22 |

From: Collins GS, Moons KGM, Dhiman P, et al. *BMJ* 2024;385:e078378. doi:10.1136/bmj-2023-078378

[1] D=items relevant only to the development of a prediction model; E=items relating solely to the evaluation of a prediction model; D;E=items applicable to both the development and evaluation of a prediction model
[2] Separately for all model building approaches.
[3] TRIPOD-Cluster is a checklist of reporting recommendations for studies developing or validating models that explicitly account for clustering or explore heterogeneity in model performance (eg, at different hospitals or centres). Debray et al, BMJ 2023; 380: e071018 [DOI: 10.1136/bmj-2022-071018
[4] This relates to the analysis code, for example, any data cleaning, feature engineering, model building, evaluation.
[5] This relates to the code to implement the model to get estimates of risk for a new individual.
